# Should individuals be paid to participate in mass drug administration to control malaria?

**DOI:** 10.1101/2022.03.16.22272499

**Authors:** Helen Fryer, Meg Peyton-Jones

**Affiliations:** Big Data Institute, University of Oxford, Old Road Campus, OX3 7LF; Wellcome Centre for Cell Biology, University of Edinburgh, Michael Swann Building, Max Born Crescent, Edinburgh, EH9 3BF

## Abstract

Mass drug administration (MDA) is a malaria control strategy in which antimalarial drugs are offered to a whole community. Although MDA can potentially clear malaria from a community, it is not routinely used in eradication efforts because of ethical concerns and past failures to achieve lasting elimination. One potential means to improving the outcome of MDA is to incentivize individuals to participate, for example, through monetary payments. In this study our aim is to inform the decision to use MDA to eradicate malaria and explore whether individuals should be incentivized to participate. Through the lens of a mathematical model, we clarify how the costs and benefits of MDA are context-dependent. We highlight that in a community experiencing a good improvement in clinical case management – relative to the prevalence of malaria in the community – the addition of MDA can catalyze stable elimination. However, participation rates are critical and individuals who avoid every round of treatment can prevent elimination. We explore how, in this scenario, individual incentives could change the cost-benefit breakdown, measured at three levels – personal, local community and wider community.

## Introduction

Malaria kills over 400 thousand people annually [1], making it one of the world’s most deadly infectious diseases. Most malaria endemic countries have demonstrated that with financial backing they can scale up their control programs and significantly reduce the disease burden [2]. The result is that the global death toll from malaria has halved since the beginning of the 21st century [1]. These gains have been made principally through widespread deployment of long-lasting insecticidal nets [3, 4], indoor residual spraying of insecticides [5, 6], rapid diagnostic tests and treatment with highly effective artemisinin combination therapies (ACTs) [7]. These and other interventions will need to be maintained and scaled up further to achieve the World Health Organization goal of reducing the annual death toll from malaria between 2016 and 2030 by 90% and eliminating the infection in over 35 endemic countries [8].

Malaria vaccine research has made significant recent advances [9], however alternative control strategies offer potential benefits that should be critically assessed. Mass drug administration (MDA) is a control strategy in which an entire community is offered antimalarial drug treatment [10]. As natural immunity to malaria is acquired gradually with repeated exposure to infection, there is progressive protection with age against the symptoms of malaria [11]. Asymptomatic malaria infections are common in endemic regions and are understood to play an important role in sustaining the spread of malaria [18]. Mass drug administration seeks to eliminate the human stage of malaria entirely and therefore has the potential to significantly reduce or clear the targeted population of asymptomatic, as well as, symptomatic infections. Mathematical models have been used to explore the potential of MDA in malaria elimination [12-15]. They have revealed that in certain contexts, MDA can rapidly clear a community of malaria. Furthermore, in some contexts, future outbreaks can be prevented by simultaneously improving other health care provisions, including the rapid testing and treatment of clinical cases.

Despite the theoretical potential of MDA and recent interest in its potential [16-18], it is not routinely used as a tool in malaria elimination. There are multiple reasons for this. There are clear ethical considerations [19] to encouraging healthy individuals to take antimalarial drugs that can cause side effects. This is particularly true when there is a risk that the intervention would not halt transmission and that any reduction in disease prevalence would be short-lived. Indeed, historically, MDA programs have had limited success in achieving a lasting, malaria free status [10, 20]. Plausible explanations for these failings include low participation and/or drug adherence; the use of poor efficacy drugs; insufficient improvement to clinical case management following MDA and a lack of effective measures to achieve elimination in adjoining regions from which individuals may travel.

Participation levels can affect the success of MDA [13, 14]. Individuals who refuse to participate can thus contribute to the failure of the intervention at the community level and render the costs to participating individuals (e.g. the drug side effects) taken in vain. For this reason, the issue of personal choice and coercion [19] in regards to participation in MDA should be carefully evaluated. Individual incentives such as monetary payment are one type of coercive action that can be effective [21]. Although payments for taking drugs are considered by some to be unethical, others argue that payments are only unethical if they are too low to compensate for the risk to the individual [22]. However, in the context of MDA, it could also be argued that it is unethical to allow some individuals to jeopardise the success of a campaign that affords a risk to others and which, if successful, would have serious health benefits to large numbers of individuals. Health policy makers are in the difficult position of having to weigh up the costs and benefits of MDA [19]. Not only do these costs and benefits act at different levels (e.g. the individual and the community), but they are context specific. The aim of the present study is to inform decisions on MDA. Specifically, we will address the following questions.

- What are the costs and benefits of mass drug administration and at what levels do they act - e.g. the personal level or at the level of the community?
- Does MDA have any benefit above just improving clinical case management?
- How is the outcome of MDA dependent upon participation rate and other factors, such as the provision of clinical case management and the relative isolation of the community from other infected communities?
- Should individuals be incentivised to participate in MDA?

By adapting a simple mathematical model of the impact of drug treatment on the spread malaria, we explore the outcome of MDA by clarifying the circumstances under which it would be expected to 1) facilitate elimination, 2) speed up elimination and 3) fail to achieve elimination in a local community. In addition, we will define the stability of the achieved elimination state to understand how robust it would be to the travel of infected individuals into the community. We will further evaluate the circumstances under which participation levels are critical to the outcomes of MDA to inform the decision to incentivise participation.

## Results

### Individual-level costs and community-level benefits

We begin this study by listing costs and benefits associated with MDA. For pragmatic reasons, any MDA program would most likely be implemented by a limited number of teams of health workers that move from one local community to another. For this reason, we have centered this work on characterizing the costs and benefits associated with an MDA program [19] that is applied to a local community. However, we note that the costs and benefits can act at multiple levels – the individual, the local community and the wider community (see Figure 1).

**Figure 1.**
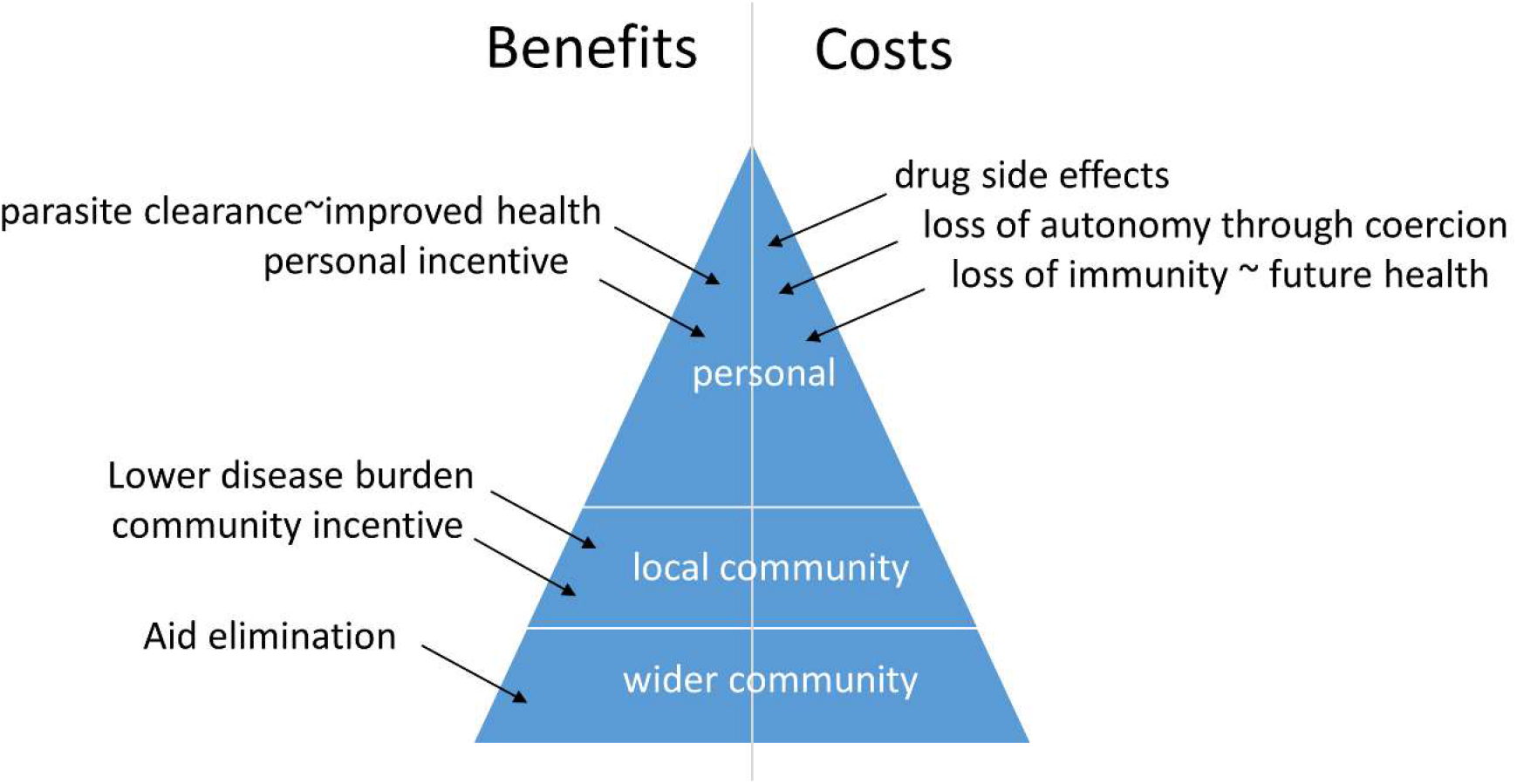
Costs and benefits of mass drug administration. Mass drug administration is associated with costs and benefits that act at different levels –the personal level, the local community and the wider community.

By considering the costs and benefits acting at these different levels, it becomes clear that participating individuals bear measurable costs, whereas the benefits of a successful MDA program can be observed amongst all levels, including the local and wider communities. Costs born by individuals include the side effects of drugs and the potential loss of immunity that could impact future health. In the event that individuals are incentivised to participate, the cost also includes a loss of autonomy. Personal benefits include improved health through parasite clearance – if the individual happens to be infected – and the receipt of a personal incentive, such as money, in circumstances when these are offered. Community-level benefits include community incentives, if offered. Other benefits (to the individual, local community and wider community) are dependent upon whether or not MDA successfully eliminates malaria from the community. In the event that sustained elimination is achieved then there is a significant future health benefit (disease absence) to the individual and the local community and this also contributes to elimination across a wider community. However, in the event that MDA does not achieve sustained elimination then the health benefit to the individual and the community is only temporary and an associated reduction in immunity could leave individuals in the community at risk of more complicated disease. This heuristic review highlights how the costs and benefits of MDA depend upon the expected outcome. Later in this study we thus seek to clarify how the outcome of MDA is context specific and therefore how the cost-benefit balance can potentially be affected by personal incentives.

### Elimination is not always achievable by improving clinical case management

It has been demonstrated both theoretically and empirically [7, 13] that improving the management of clinical cases, not only reduces mortality due to malaria, but also reduces the spread of malaria amongst a population. Arguably, if improving clinical case management is sufficient to facilitate malaria elimination, improving such a provision is favourable to MDA, as it avoids the treatment of malaria-free individuals. Before evaluating the impact of MDA in detail, our aim is to first clarify the circumstances under which clinical case management, alone, can lead to elimination.

An existing transmission model of malaria [12, 13] was adapted to address the study questions in a transparent manner. The full model, explained in detail in the Methods section, characterises the spread of malaria amongst a human population. Infected individuals progress through three different infection stages (Figure 2). These are first, the asymptomatic stage, second the symptomatic stage and, third, the infectious stage (which is also symptomatic). Infection is followed by recovery. An adjustable fraction of such recovery events lead to clinically immunity causing further infections being asymptomatic. The possibility that clinical immunity can lead to a reduction in infectiousness is also modelled [23]. A chosen proportion of all symptomatic individuals receive clinical treatment, which increases the recovery rate. In addition, a chosen proportion of all individuals receive mass drug administration, which increases the recovery rate of both symptomatic and asymptomatic infections. There is host turnover and fluctuations in the probability of transmission to implicitly account for seasonal changes in the prevalence of the mosquito over the course of each year.

**Figure 2.**
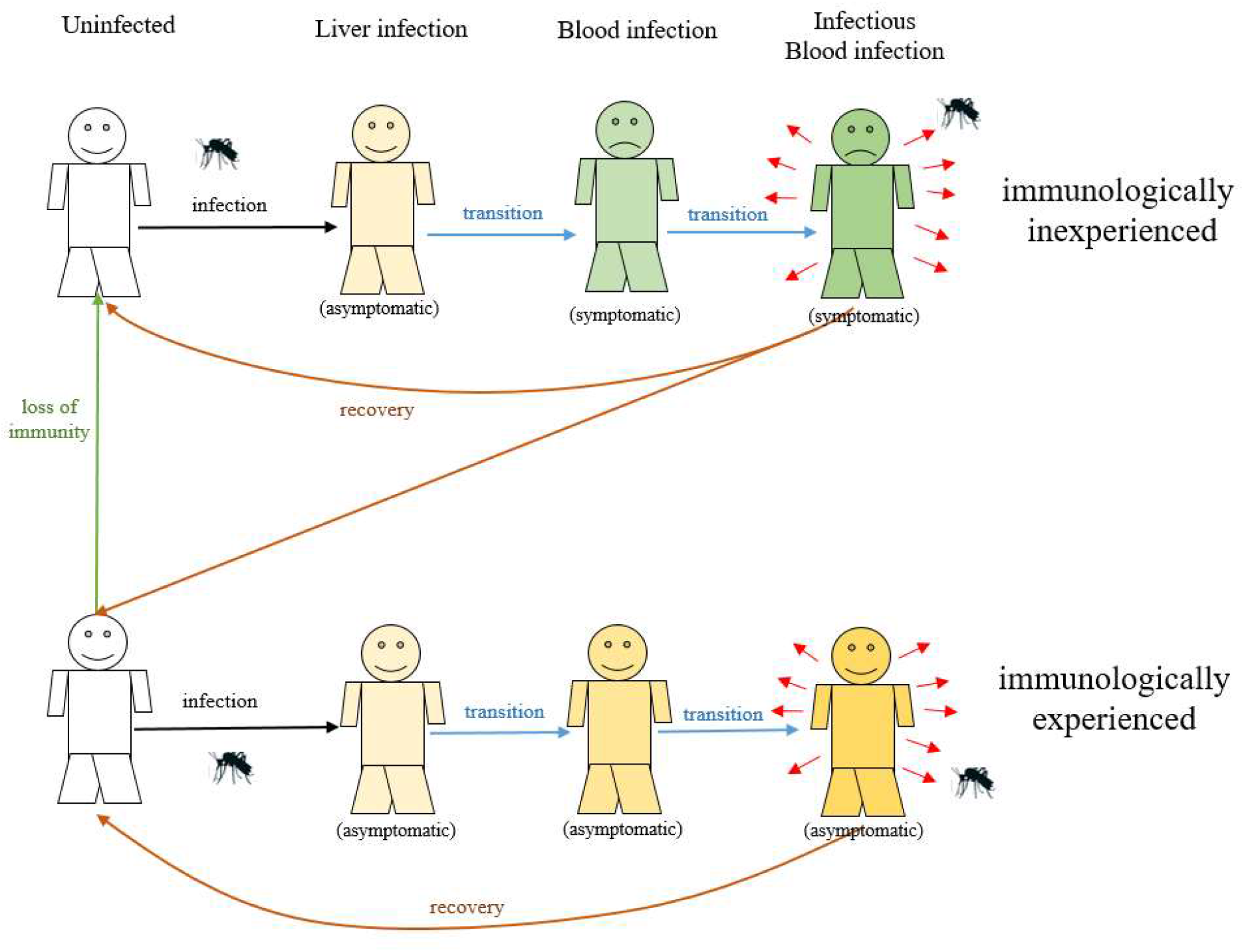
How natural infection is modelled. This figure illustrates how natural infection is represented in the compartmental model. The full model (see Supplementary Figure 3 and Supplementary Text 1) also includes host turnover and the impact of clinical treatment and mass drug administration.

In Figure 3 we use the model to explore the impact of clinical treatment on the spread of malaria. In Figure 3a), prior to time 0, a malaria epidemic with a mean malaria prevalence of 30% is assumed to be ongoing amongst a population of 1000 individuals. Beyond time 0, 40% of individuals are assumed to receive clinical treatment upon symptomatic infection (black line) resulting in a gradual reduction in the prevalence of malaria, such that the infection is eliminated (defined as there being fewer than 1 infected individual remaining), 15 years later. This highlights that, under in certain circumstances, improvements to the provision of clinical treatment can lead to elimination within a community. Provided this provision is maintained, malaria be expected to remain absent from the community. However, whether elimination is predicted, and how long it would be expected to take is dependent upon a number of factors. Treatment coverage is clearly critical. In the same epidemiological scenario, 60% coverage (blue line) yields elimination in approximately a third of the time (5 years), but 20% coverage (red line) is insufficient for elimination.

**Figure 3.**
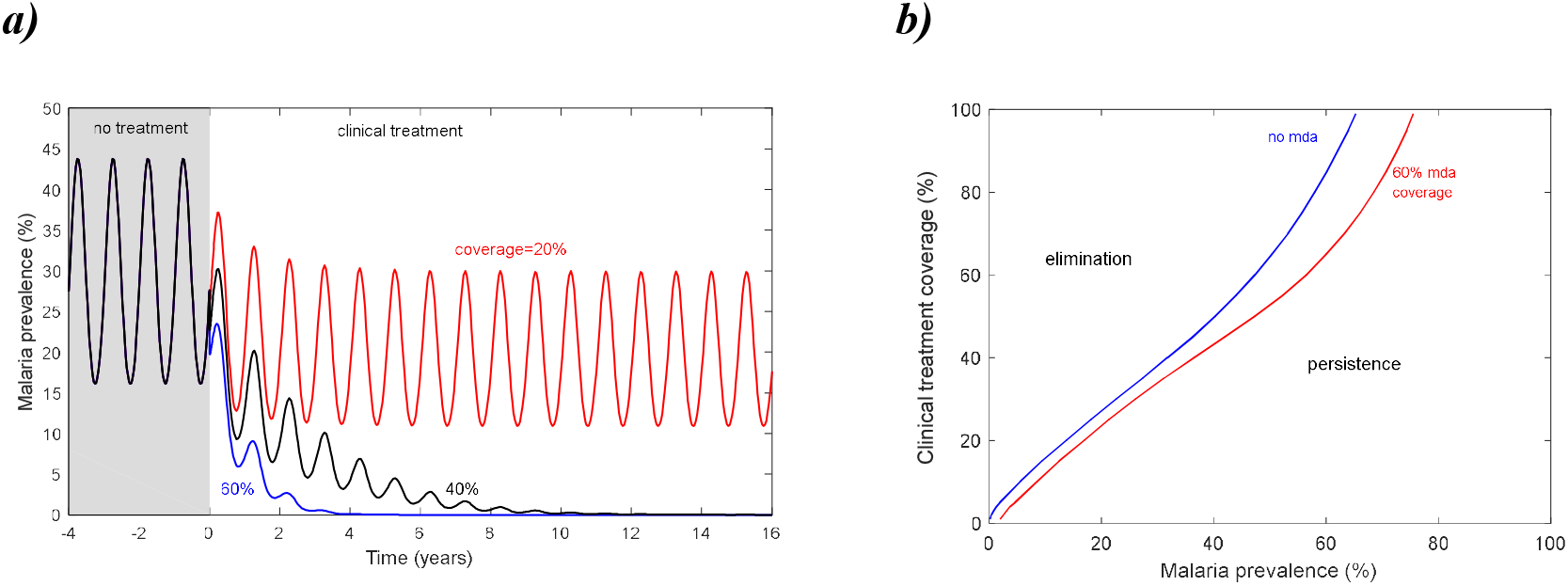
The impact of clinical case treatment on malaria dynamics. This figure explores how the prevalence of malaria in a community is affected by the treatment of clinical cases. In a) prior to time 0, a malaria epidemic is ongoing in the absence of clinic treatment and the prevalence of malaria has settled into a periodic cycle that fluctuates around a constant mean of 30% prevalence. At time 0, a given proportion of symptomatic infections receive clinical treatment. In red the treatment coverage (20%) is sufficiently low that malaria persists indefinitely and settles into a periodic cycle that fluctuates around a lower constant mean. If the treatment coverage is beyond a critical threshold, the prevalence of malaria continuously declines, such that elimination is ultimately predicted. The expected time to elimination is dependent upon the coverage – 15 years when coverage is 40% (black line) and 5 years when coverage is 60% (blue line). In b) the threshold clinical treatment coverage required for elimination is plotted (blue line) against the average malaria prevalence. If MDA with a coverage of 60% is additionally applied, the clinical treatment coverage required for stable elimination is modestly reduced (red line).

The transmission rate of malaria is also an important factor in determining the elimination metrics. The prevalence of malaria in the population is dependent upon the transmission rate and is therefore an indicator of the control effort requires to achieve elimination. Figure 3b demonstrates how the coverage of clinical treatment required to achieve elimination is larger if the prevalence of malaria in the population is larger. It also highlights that when the prevalence of malaria is above a certain threshold, elimination is not possible, even with a clinical treatment coverage of 100%.

The existence of this threshold is due to the fact that in areas where the prevalence of malaria is higher, there is a greater degree of acquired immunity in the population and so the portion of infections that are asymptomatic is higher. Understanding the biology of asymptomatic infections in regards to their infectiousness and infectious period is important for evaluating when elimination is expected. If asymptomatic infections, on average, result in at least one more asymptomatic infection over their infectious period, malaria elimination cannot be achieved through clinical treatment alone. There is currently no consensus on the relative infectiousness of symptomatic and asymptomatic blood stage infections [23]. In Figure 3. they are assumed to be equally infectious, but if asymptomatic infections are less infectious than symptomatic infections the maximum prevalence of malaria that can be eliminated through clinical treatment is higher (Supplementary Figure 1). It is also noteworthy that the threshold criteria is dependent upon the rate that immunity is lost and acquired (for which some degree of uncertainty remains) and thus the proportion of infections that are asymptomatic. If a greater proportion of infections are asymptomatic then then the maximum prevalence of malaria that can be eliminated through clinical treatment is lower (Supplementary Figure 2).

### High participation MDA can ‘catalyse’ stable elimination

The fact that improvements to clinical case management are not always sufficient to stop the spread of malaria in a community points to the need for additional interventions to achieve elimination. Alternative control measures include the deployment of long-lasting insecticidal nets and indoor residual spraying of insecticides [5, 6]. However, if these combined measures are already in place and insufficient to achieve elimination, others must be sought. MDA offers a potential means to improve the contribution that drug therapy can make to the goal of eliminating malaria. Towards evaluating whether individuals should be incentivised to participate in MDA, here our aim is to clarify the context under which MDA can be a useful tool in malaria elimination. Part of this aim is to clarify how participation rates contribute to the outcome of MDA.

In Figure 4 we compare the impact on an ongoing malaria epidemic of two uses of drug treatment: 1) the treatment of a chosen proportion of clinical cases, and 2) the treatment of a chosen proportion of clinical cases in addition to mass drug administration. Mass drug administration is assumed to consist of three rounds of drug treatment (therapy and piperaquine) that each begin 30 days apart [7]. In each round, a chosen percentage of the population is randomly selected to receive therapy. ACT remains effective for 3 days and piperaquine remains effective for 25 days [24]. Thus, there is a five day period between rounds when the drug is no longer effective.

**Figure 4.**
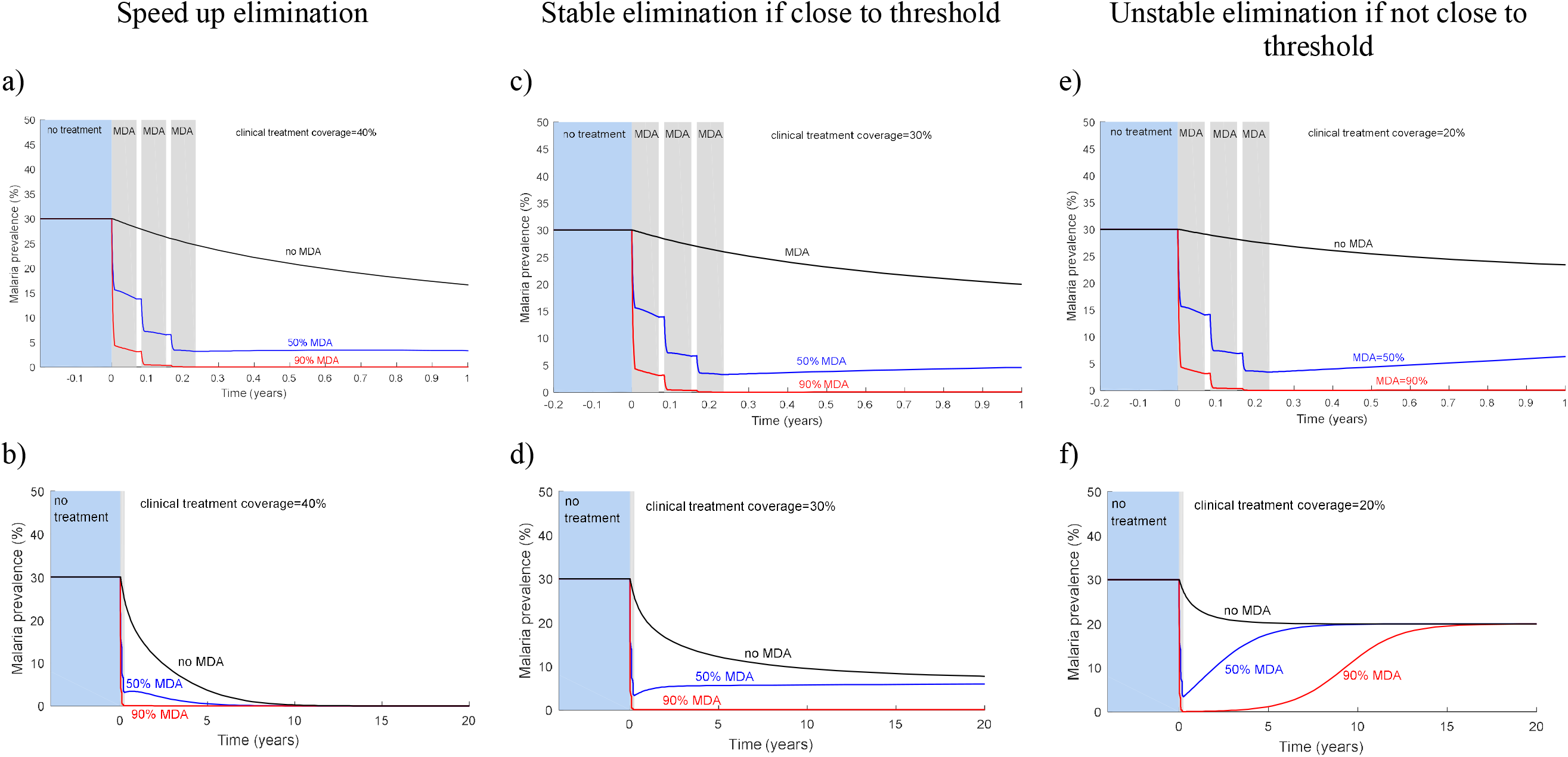
The combined impact of clinical treatment and MDA on malaria dynamics. This figure explores how the prevalence of malaria in a community is affected by antimalarial drug interventions. In each simulation, seasonality has been removed from the model to illustrate the principal dynamics, which are not affected by this simplification. Three different clinical treatment scenarios are explored. In each scenario, prior to time 0, a malaria epidemic with a prevalence of 30% is modelled in the absence of any drug treatment. At time 0, three interventions are modelled. One in which clinical treatment at a chosen coverage begins (black line) and two in which three rounds of MDA, starting a month apart, are additionally applied at either a low coverage (50%; blue line) or a high coverage (90%; red line). For each clinical treatment scenario, the malaria dynamics are revealed over a short (panels a, c and e) timeframe and over long timeframe (panels b, d and f). We define elimination as there being fewer than 1 infected individual remaining in this population of 1000. In panels a and b) the coverage of clinical case treatment (40%) is sufficient to achieve elimination, but it takes 11.3 years. The addition of mass drug administration dramatically speeds up elimination which is achieved after 7.5 years with 50% MDA coverage or 2 months with 90% coverage. In panels c) and d) the coverage of clinical case treatment is just short of sufficient to achieve elimination (30%). The addition of MDA is able to facilitate elimination, provided the MDA coverage is high enough. Furthermore, elimination is stable once the MDA course has finished. In panels e) and f) the coverage of clinical case treatment (20%) is markedly too low to achieve elimination. The addition of MDA is sufficient to result in elimination, provided the MDA coverage is high enough. However, irrespective of MDA coverage, malaria prevalence resurges once MDA ceases to the level expected without the additional MDA.

Three different scenarios are explored to highlight the expected impact of MDA. In the first scenario (Figure 4a-b), the coverage of clinical cases alone is sufficient to achieve elimination (defined as previously), but it takes several years. The addition of mass drug administration can dramatically speed up elimination. Assuming that clinical treatment continues at the same coverage once MDA stops, elimination is stable. In real terms this means that ongoing transmission would be halted and that even if a few infected individuals were to travel into the community from elsewhere, they would not be able to spark a new epidemic in the community. In another scenario (Figure 4e-f), the coverage of clinical treatment is markedly too low to achieve elimination. If mass drug administration is additionally undertaken, elimination is predicted, provided the coverage is high enough. However, elimination is not stable once the MDA programme stops. In real terms this means that if some infected individuals were to travel to this community from elsewhere, malaria would be expected to spread amongst the population once more. There also an intermediate scenario for which the dynamics are, arguably, counterintuitive (Figure 4c-d). Here, the coverage of clinical treatment is just short of sufficient to achieve elimination. If mass drug administration is additionally undertaken, provided the coverage is high enough, elimination occurs and remains stable once MDA stops. In real terms, therefore, ongoing transmission would be halted and infected travellers to the community would not be able to spark a resurgence. This scenario highlights that it is more difficult to achieve malaria elimination though clinical treatment, than it is to maintain it. Mass drug administration can act as a ‘catalyst’, tipping the epidemiological system into a state where control is easier. This is highlighted by the blue line in Figure 3b which shows that MDA lowers the clinical treatment coverage threshold required for elimination.

In mathematical terms, this dynamical system is known as ‘bistable’. It has two steady states: an infected one and an uninfected one; and the state that prevails depends upon the starting conditions. All other things being equal, if the starting conditions are close to one state, that state will result. The model prediction that malaria elimination is more difficult to achieve than it is to sustain can be attributed to the principle that as malaria prevalence increases, there is more acquired immunity in the population and a greater proportion of infections are asymptomatic [25, 26]. A smaller proportion of infected individuals thus seek clinical treatment (which shortens the infectiousness period) and this promotes persistence. By contrast, when the community is close to elimination, immunity is sparse and most infections are symptomatic. A higher proportion of infected individuals seek clinical treatment and this promotes elimination. The criteria for bistability has been explored elsewhere and demonstrated to occur when, on average, immunologically experienced infected individuals generate more new infections than immunologically inexperienced infected individuals [25]. This concept of the ‘stickiness’ of malaria elimination is not restricted to modelling theory. Although malaria has resurged in most countries that have tried to eliminate it, a review of resurgence following (at least temporary) elimination, records only four such failures out of 50 successful programmes [27].

The outcome of MDA is not only dependent upon the coverage of clinical case treatment. Malaria prevalence and the coverage of mass drug administration are both important. The overlap between participants in each round of MDA is furthermore significant [14]; if there is a lack of independence in the people who participate in each round (i.e. certain people fail to participate in all three rounds), the critical participation rate required for elimination is higher than under the assumption of independent sampling. With a view to evaluating whether individuals should be incentivised to participate in MDA, Table 1 summarises how the outcome of MDA depends upon the context and upon participation rates.

**Table 1.** Under what circumstances would MDA lead to localised elimination of malaria and how is this impacted by MDA participation rates? This table explores how the expected outcome of MDA in a local community is not only dependent upon participation rates, but also on other factors. The degree of isolation from another infected community and the level at which clinical case management is scaled up (relative to the prevalence of malaria in the community) is also critical for determining whether elimination is expected, how long elimination takes, how stable this state of elimination is and how resurgence would take.

| Isolation from infected community | Improvements to clinical treatment relative to malaria prevalence | MDA participation |  |  |
| --- | --- | --- | --- | --- |
|  |  | Low | Intermediate | High |
| Low | -- | No | No | No |
| Intermediate | Poor | No | No | No |
| High | Poor | No | No | Possible (unstable) |
| Intermediate/High | Good | No | Possible (stable) | Yes (stable) |
| Intermediate/High | Very good | Yes (stable) | Yes (stable) | Yes (stable) |

slow fast Time to elimination

Although we have not explicitly modelled the movement of infected individuals into and out of the community, the impact of the relative degree to which a community mixes with other infected communities, can be inferred from understanding the stability of elimination once MDA is stopped. In summary, MDA participation rates are not expected to change the long-term outcome of MDA (persistence) when improvements to clinical case management (relative to the prevalence of malaria) are poor or when the community mixes readily with other infected communities that are not subject to the same intervention. MDA participation rates would also not change the ultimate outcome (elimination) when improvements to clinical case management are very good and the community is at least partially isolated from other infected communities, though they would change the timeframe of elimination. However, in a similarly isolated community, when improvements to clinical case management are good, MDA participation is critical to the outcome. Stable elimination is expected beyond a certain critical threshold, but persistence is expected below it.

### Individual incentives could significantly change the cost-benefit breakdown

We sought to explore whether individuals should be incentivised to participate in MDA, for example through monetary payments. As we have shown above, the proportion of individuals who participate in a community MDA programme can (depending upon the context) be critical to its success in halting transmission. For these reasons one might argue that MDA programs should only be undertaken in situations where there is a good chance that participation levels will be beyond the required critical threshold. Experience from the Thai-Myanmar border has demonstrated that with good community engagement and community-level incentives that participation rates that are sufficient to halt transmission can be achieved. However, localised elimination has not been achieved in all communities on the Thai-Myanmar border that have undertaken MDA. One potential explanation for this is that participation rates in some communities were too low. Individual incentives, such as momentary payments are a plausible means to increasing participation in MDA. However, they are often discounted on ethical grounds because payment levels that are high enough to be persuasive impose some loss of autonomy on the individual in regards to the individuals’ decision to participate. Acknowledging this argument, we nevertheless believe that individual incentives for participation should not be discounted without a clear evaluation of the impact that they could have on the full cost-benefit breakdown.

In Figure 5 we explore how the costs and benefits of MDA would be expected to change according to whether individuals are incentivised to participate. We focus on what is arguably the most compelling scenario for incentives: clinical treatment alone is assumed to be just short of sufficient to achieve elimination; without incentives (Figure 5a), participation in the additional MDA is too low to achieve elimination, but with incentives, it is high enough to achieve stable elimination (Figure 5b). When there are no incentives, uninfected individuals who participate do not receive any measurable (personal) benefits and bear the burden of side effects in pursuit of very limited benefits to either the local or the wider community. When personal incentives are used, however, there is a significant change to the cost-benefit breakdown. Participating individuals receive the personal benefit of both the incentive (e.g. the payment) and a possible improvement to their future health through the absence of future malaria infections. In addition, stable elimination affords great benefit to the local community (including family members), particularly children, whose lives may be saved. There is also a further contribution to eradication efforts amongst the wider community through the reduced spread to other regions. Policy makers must evaluate whether these multi-level benefits equate to an ethical ‘advantage’ to personal incentives and outweigh the ethical ‘cost’ associated with a loss of autonomy through this form of coercion. Clearly, the cost-benefit breakdown will vary according to context. For example if clinical treatment coverage is markedly too low to achieve elimination alone and the community mixes regularly with other infected communities, individual incentives offer no benefit to individual future health. They also offer no measurable benefits to the local community or the wider community. In such a scenario, the personal costs (side effects and loss of autonomy) are countered only by the value of the incentive and a possible improvement to health if the individual happens to be infected.

**Figure 5.**
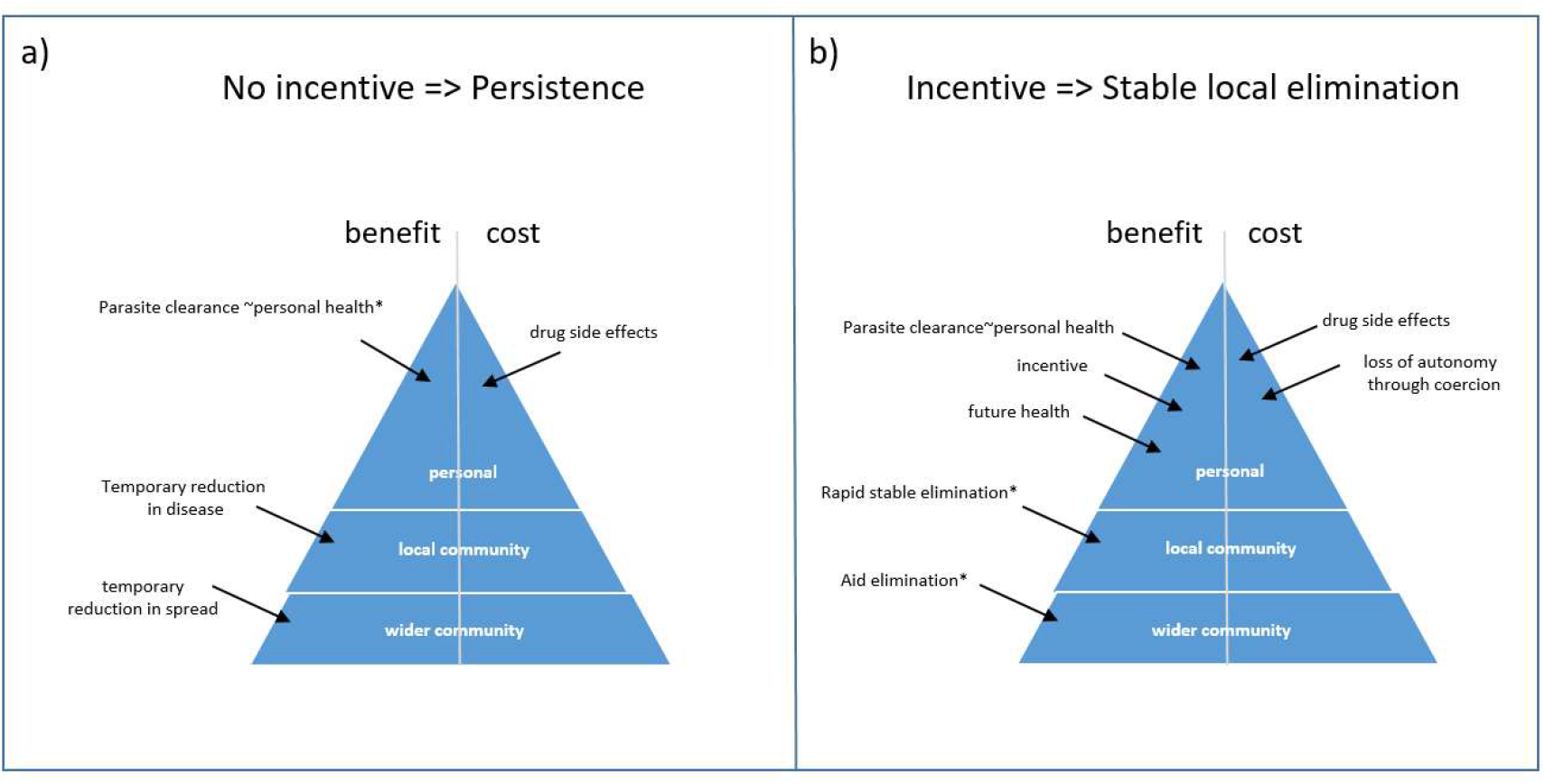
A comparison of the expected costs and benefits with and without individual incentives. Here we consider an epidemiological scenario in which clinical treatment is just short of sufficient to achieve elimination. We explore how individual incentives could impact the costs and benefits of also including MDA. In a) there are no incentives and participation in MDA is assumed to also be too low to achieve elimination. In b) individuals are incentivised and participation in MDA is assumed to be sufficiently high to achieve stable elimination.

## Discussion

The aim of this study was to inform decisions into the use of MDA to aid malaria eradication and to explore whether individuals should be incentivized to participate in MDA. For pragmatic reasons, any MDA program would most likely be implemented by a limited number of teams of health workers that move from one local community to another. For this reason, we have centered this work on characterizing the costs and benefits of a community MDA program. To explore how the outcome (i.e. the benefit) of MDA is dependent upon context, we have adapted an existing mathematical model of the impact of drug treatment on the transmission of malaria to offer maximum transparency. Using this model, we highlight that the prevalence of malaria in the community is an important indicator of the effort required to achieve elimination through any intervention, including MDA. Although improvements to the treatment coverage of clinical cases can be sufficient to achieve elimination when the prevalence of malaria is below a critical threshold, beyond it, other interventions are required. MDA is one option as it can catalyze stable elimination in communities where clinical treatment is also significantly up-scaled (relative to the prevalence of malaria in the population). However, participation rates are critical to the outcome of MDA and individuals who avoid every round of treatment can prevent elimination. We have also described how, in this context, individual incentives could significantly change the cost-benefit breakdown, as measured at three levels – individual, local community and wider community. In other contexts, the argument for individual incentives for MDA participation is weaker. Notably, in communities that mix frequently with other infected communities and where clinical treatment is not simultaneously up scaled, malaria is expected to continue to transmit, irrespective of MDA participation. This reinforces the advice that targeting individual communities just with MDA is unlikely to have any lasting impact, whereas a comprehensive health program that includes clinical treatment as well as MDA and which sweeps through multiple communities across a wide region, such as a country, is expected to have a much greater chance of success.

Nevertheless, there are heterogeneities between local communities (for example, malaria prevalence and intervention uptake) that mean that if MDA were rolled out across a wide community as a series of local-community programs, the expected impact across these local communities would be varied. Policy makers must consider the ethics of using MDA in the knowledge that it will fail in some communities and whether it is appropriate to repeat it in these communities. However, it is noteworthy that even if MDA does not immediately achieve localized elimination in certain communities, the achieved reduction in the prevalence of malaria would contribute to the broader goal of achieving elimination across a wider community if efforts are sustained. There exists scope to develop models that acknowledge heterogeneities across communities to understand its impact on achieving widespread elimination.

We have intentionally focussed on the qualitative impact that participation rates can have on elimination prospects using MDA. However, multiple factors determine the elimination criteria and the expected time to elimination and their role has been quantitatively estimated with models that also account for stochastic effects, elsewhere [14, 23, 26, 27]. These include intractable factors, such as the relative susceptibility and infectiousness of symptomatic and asymptomatic infections, the population size and the rate that immunity is lost in uninfected individuals. They also include tractable factors, such as the number and spacing of rounds and the effectiveness and half-life of therapy. Any evaluation of whether individuals should be incentivised to participate in MDA must acknowledge that it may be possible to manipulate alternative factors to achieve the same outcome. For example, increasing the number of treatment rounds is one route to improving the outcome of MDA, but the number of rounds of drug therapy that healthy individuals should be encouraged to take is open to a separate ethical debate.

The evolution and spread of antimalarial resistance poses a considerable threat to the treatment and control of malaria [28]. Although we have not explicitly modelled the impact of resistance [12, 13], our principal conclusions are robust to this simplification. ACT is the first line therapy for the clinical treatment of plasmodium falciparum and has also recently been used in MDA. Resistance to ACT does not currently ameliorate the effect of the ACT, but rather, it increases the parasite clearance half-life by more than two-fold [29]. The half-life of parasite clearance impacts numerical estimations of the MDA coverage required for elimination, but does not change our qualitative account of the expected outcome of MDA and the role than individual incentives for participation could play.

The contribution that MDA could make to the evolution and spread of antimalarial resistance is also an important consideration. Models have previously confirmed that the proportion of malaria cases that are resistant to antimalarials increases with their use, whether for clinical treatment or MDA [12, 13]. This, however, does not necessarily mean that antimalarial drugs should be ruled out for use in MDA. Huge reductions in the spread of malaria have been made in recent years and antimalarial drugs, particularly ACTs, have played a significant role in this. To ignore the potential of MDA could be a missed opportunity, particularly in areas where all other control measures are insufficient for elimination. Nonetheless, in circumstances where alternative measures of malaria elimination, such as the use of vaccines and insecticide treated bed nets, are sufficient and currently underused, improving the provision of these alternative measures should be a priority as they are preferable from the point of view of retaining the efficacy of ACTs for as long as possible.

The theme of this study was to ask whether individuals should be incentivised to participate in MDA to improve eradication efforts. However, as this study has highlighted, effective clinical case management is also critical to preventing malaria resurgence once MDA finishes and in some communities can be sufficient to halt transmission without MDA [7]. The fraction of clinical cases that are treated at any given time is critical to the outcome. This fraction will depend upon the time between the onset of symptoms and the start of therapy, a characteristic that we have not modelled explicitly. Improving the provision of clinical case management is clearly key to improving the waiting time to treatment and, thus the proportion of symptomatic individuals who are treated. However, we propose that individual incentives for clinical case treatment could also be considered in certain situations, as a means to improving this metric further. Although we have modelled malaria infections as being either symptomatic or asymptomatic, in reality, individuals can present with symptoms across a spectrum, from mild to severe [30]. Individuals with mild symptoms and for whom acquiring antimalarial drugs is inconvenient, could potentially be persuaded to seek prompt treatment through individual incentives. Arguably, it is more ethical to incentivise clinical treatment than mass drug administration because even in the case of asymptomatic or mildly symptomatic infection, parasite clearance can benefit health [31].

Throughout our discussion, we have assumed that individual incentives could improve participation rates in MDA and/or clinical treatment programs. However, whether this assumption is valid requires evaluation through trials. Individual incentives could make prospective participants more wary about the side effects of treatment and even reduce participation rates. Furthermore, other methods to improve participation rates should be considered. Community-level incentives – such as ancillary care and water supply infrastructure – have been offered to communities participating in MDA in the Greater Mekong Subregion [32, 33]. When combined with strong community engagement programs they have, in some local communities, achieved levels of participated that are good enough to lead to localized elimination that has lasted for the duration of the study period [7]. In other local communities in the same region malaria has persisted. For this reason, we believe that the role of paid participation should be further explored.

## Method

### Transmission model of malaria

We adapted an existing mathematical model of the impact of drug therapy on the spread of malaria amongst a human population. This is formulated as a compartmental model (Supplementary Figure 3 and Supplementary Text 1), whereby each compartment records the number of individuals in a particular epidemiological class. In the model, the population is split according to whether individuals are participating in a mass drug administration program (*i* = 1) or not (*i* = 0). New individuals are born into an immunologically inexperienced, susceptible, state at rate *μ*. A proportion, *θ*, of these participate in MDA (*S*_0_) and the remaining proportion (1 − *θ*), do not (*S*1). All individuals in the population die at a per person rate of *μ* years^-1^.

Amongst the fraction of the population who do not participate in MDA, immunologically inexperienced individuals are infected at a rate that is proportional to the transmission function, *λ*(*t*). This function incorporates seasonal fluctuations in the probability of transmission and accounts for the fraction of individuals in the population who are infectious, as well as the relative degree of infectiousness of these individuals (equation 1. Supplementary Text 1).

The first stage of infection is an infection of the liver (*L*_0_) that is neither infectious, nor associated with clinical symptoms. Transition from this state to a symptomatic, but uninfectious, blood-stage infection occurs at a per person rate of α years-1. Upon transition, a fraction, *p*, proceed to a treated stream (*B*_0_), and the remaining fraction (1− *p*), to an untreated stream 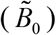. Amongst each stream, transition to a symptomatic and infectious blood stage infection occurs at rate *σ* years^-1^. The infectiousness of immunologically experienced individuals in this final stage of infection is assumed to be reduced by a factor *η*, compared to immunologically inexperienced individuals in the final stage. Recovery occurs at rate 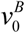 years^-1^ amongst treated individuals in the uninfectious blood stage of infection, 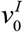 years^-1^ amongst treated individuals in the infectious stage and 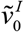 years^-1^ amongst untreated individuals in the infectious stage. A fraction *z* of all recovered individuals transition to an immunologically experienced, uninfected state 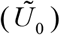 and the remainder transition back to the immunologically inexperienced, uninfected state. Recovered individuals acquire infections at the same rate as susceptible individuals and transition through the three stages at the same rates. As they are immunologically experienced, all stages are asymptomatic and only infectious stage individuals recover – always to an immunologically experienced state.

Amongst the fraction of the population who participate in MDA, the dynamics are equivalent, with the following exceptions. There is no distinction between treated and untreated individuals; all individuals are treated and therefore there is recovery from all infected classes. The rate of recovery of immunologically inexperienced individuals in the liver stage, uninfectious blood stage and infectious blood stage is 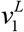 years^-1^, 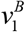 years^-1^ and 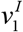 years^-1^, respectively. The equivalent rates amongst immunologically experienced individuals are 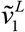 years^-1^, 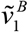 years^-1^ and 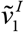 years^-1^, respectively. All model parameters and variables are listed in Supplementary Tables 1 and 2).

## Data Availability

THere are no data in this study

---

**Supplementary Figure 1.**
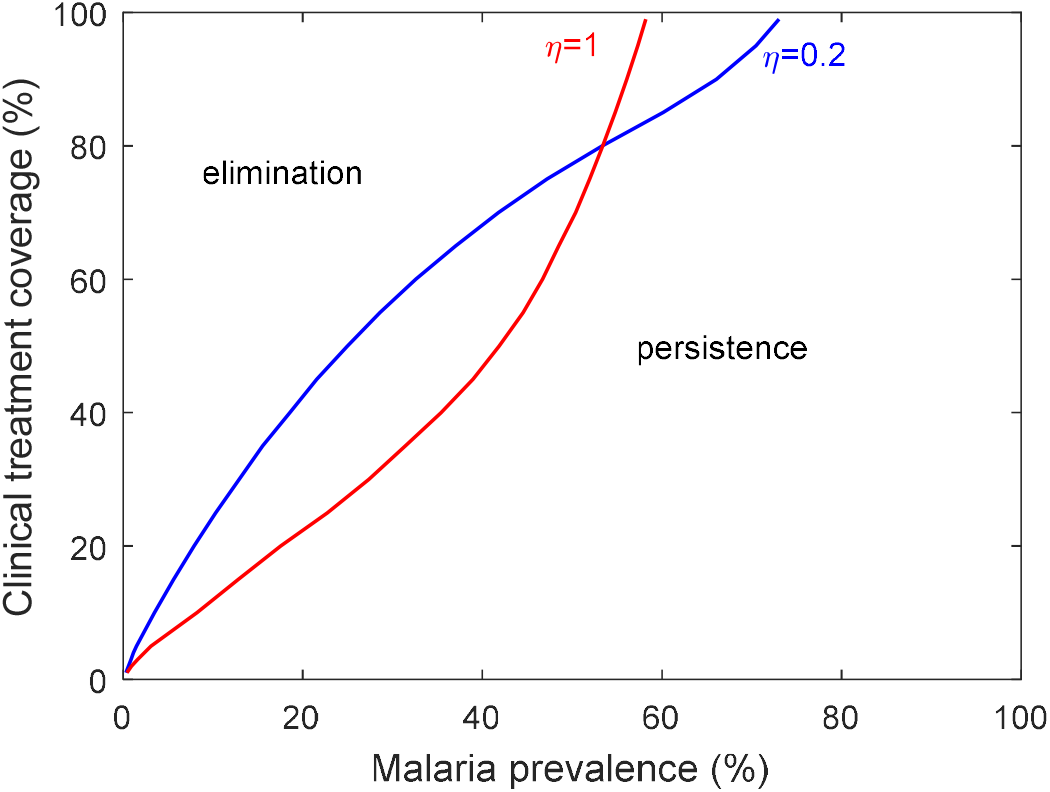
The infectiousness of clinically immune infected individuals affects the clinical treatment threshold. This figure reveals how the relative infectiousness of immunologically experienced and immunologically inexperienced individuals impacts the threshold clinical treatment required for elimination (in the absence of MDA). In blue immunologically experienced infected individuals are one fifth as infection as immunologically inexperienced individuals (η=0.2 years-1) and in red the two host classes are equally as infectious (η=1 years-1).

**Supplementary Figure 2.**
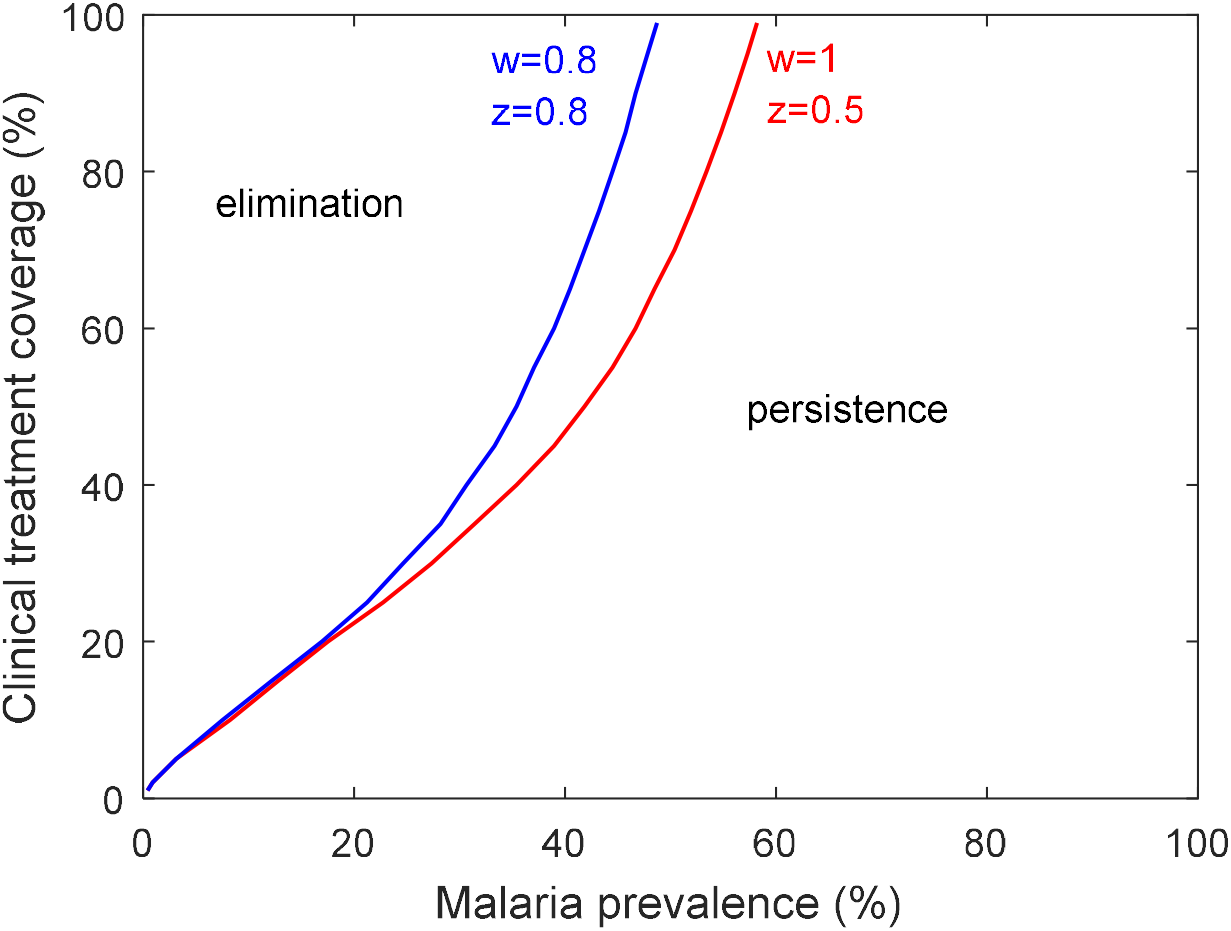
The rate that immunity is acquired and lost affects the clinical treatment threshold. This figure compares the clinical treatment threshold (in the absence of MDA) required for elimination for different rates that immunity is lost and acquired in uninfected individuals and therefore the portion of the infected population that have acquired immunity. In red immunity is lost at rate w=1 years^-1^ and 50% of recovered infections (z=0.5) yield immunity. In blue immunity is lost at a slower rate (w=0.8 years-1) and a larger proportion of infections result in immunity (z=0.8).

**Supplementary Figure 3.**
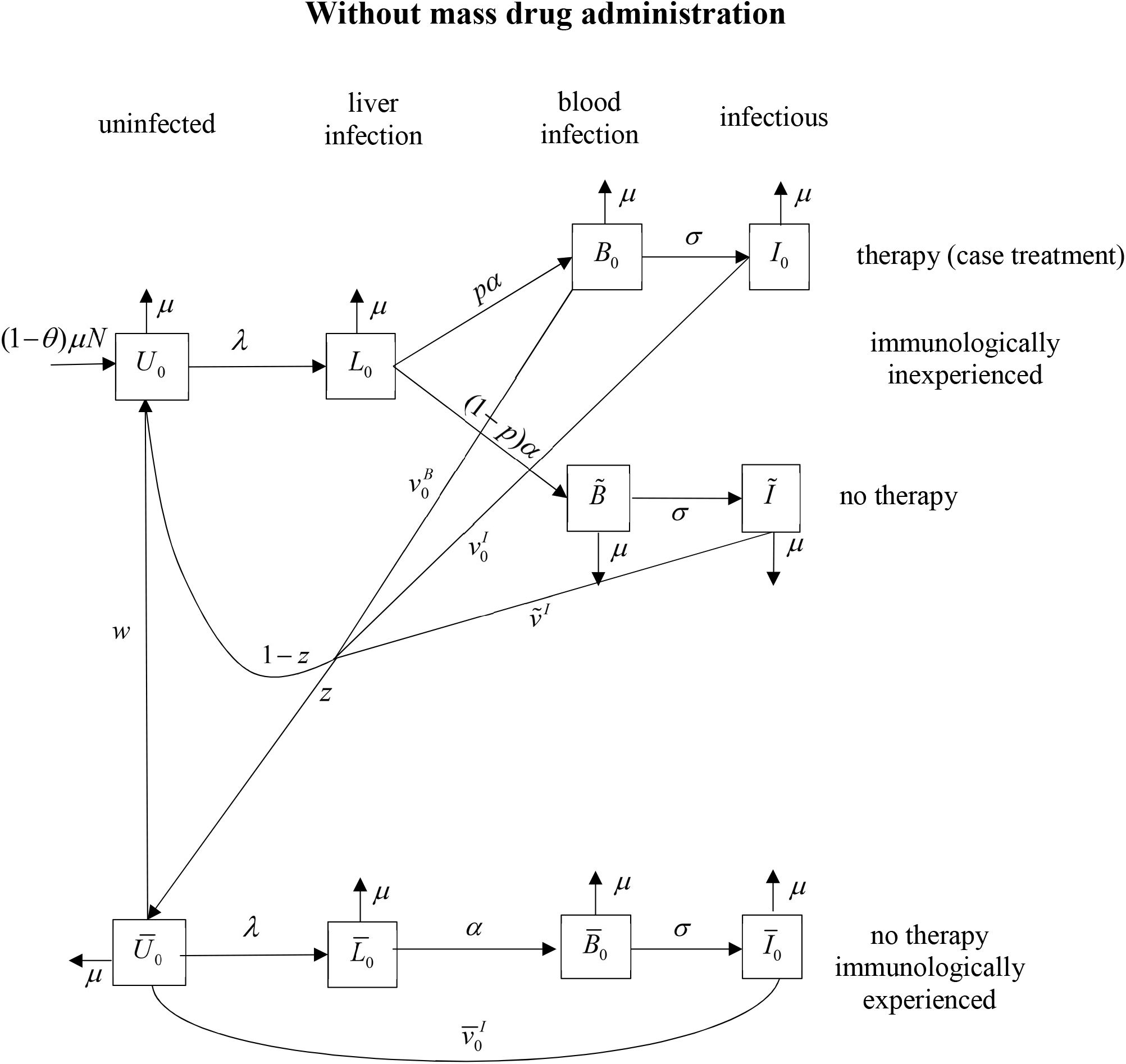
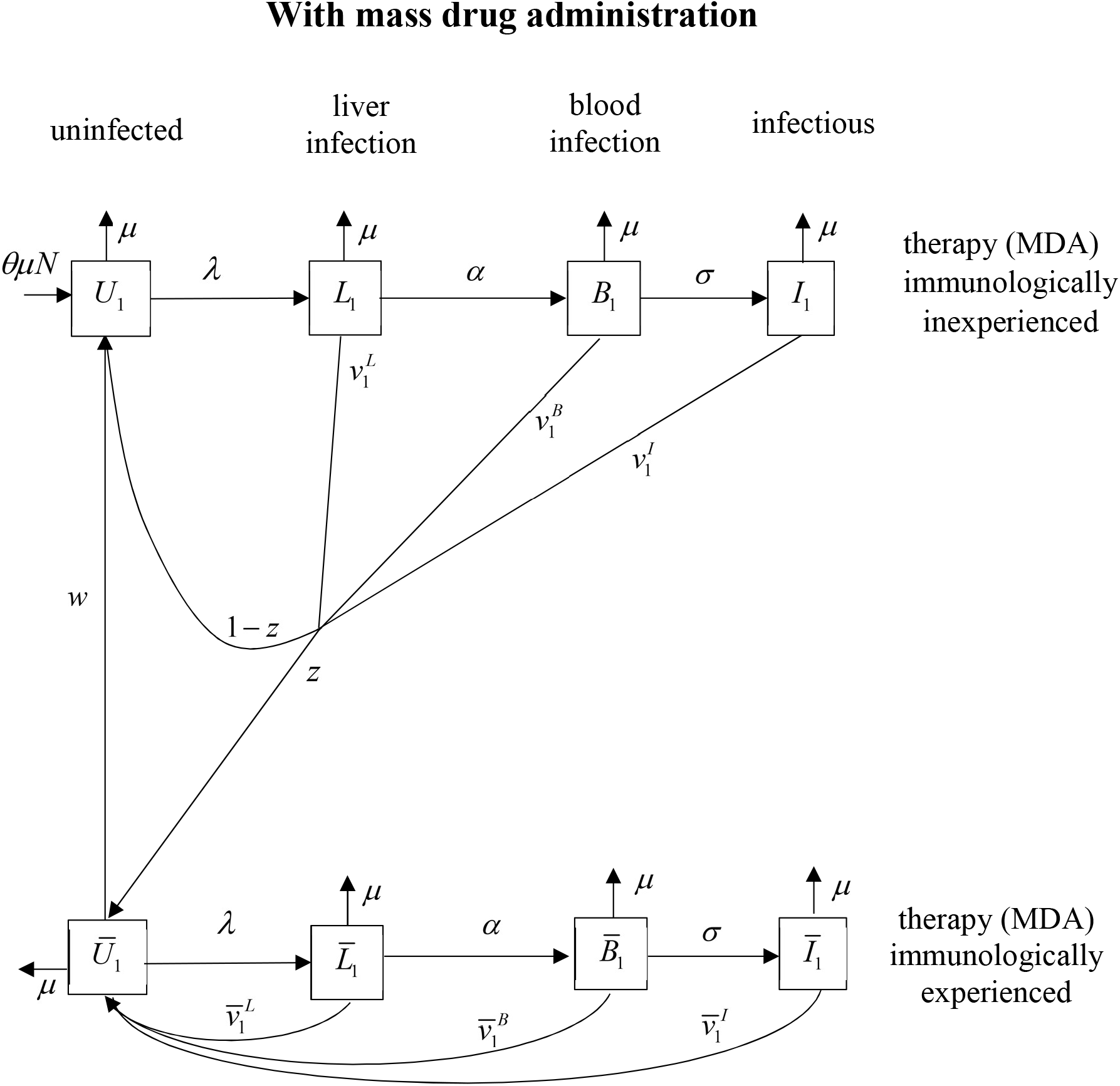
A mathematical model of the impact of drug treatment on the spread of malaria. This figure illustrates that in the mathematical model, the population is split according to whether or not individuals will receive MDA. Within each section of the population, there is the progression of disease through different stages and the acquisition of immunity that can lead to asymptomatic infections. Amongst individuals who do not receive MDA a certain fraction of those who progress to symptomatic disease receive clinical treatment. MDA increases the recovery rate at all infection stages and clinical treatment increases the recovery rate of just the two latter (blood) stages of infection. The whole population undergoes host turnover.

## Supplementary Text 1. Model equations

### Transmission function

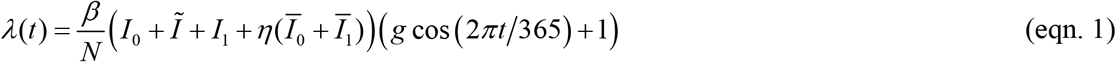

### Ordinary differential equations

Without mass drug administration

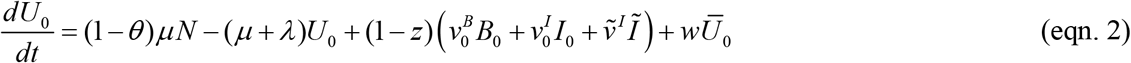

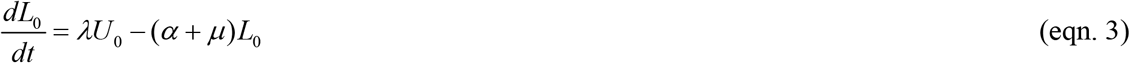

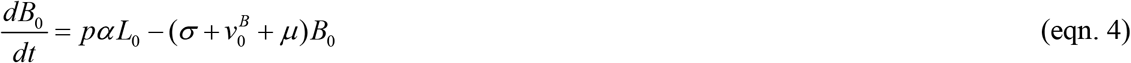

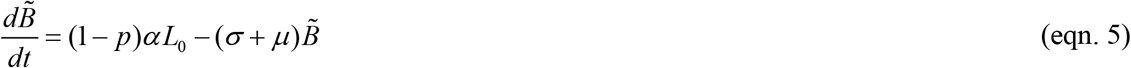

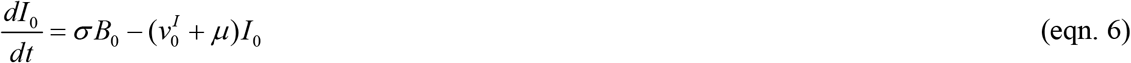

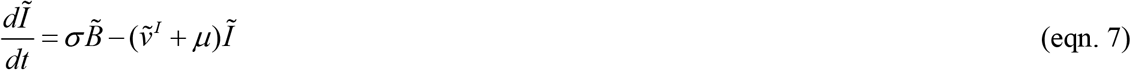

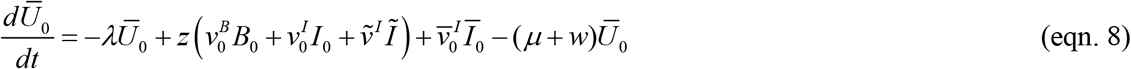

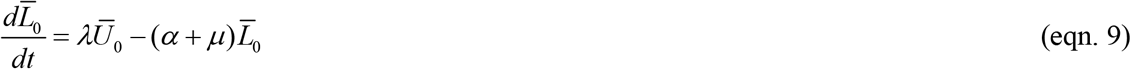

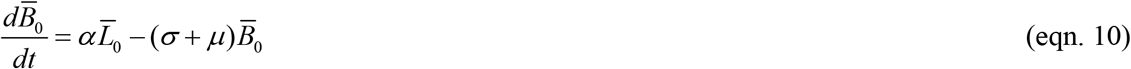

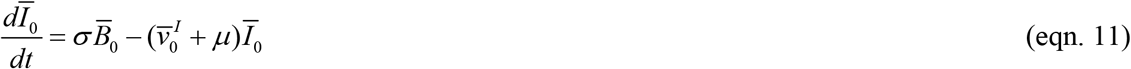

With mass drug administration

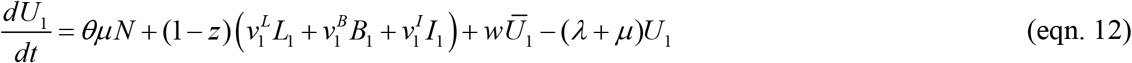

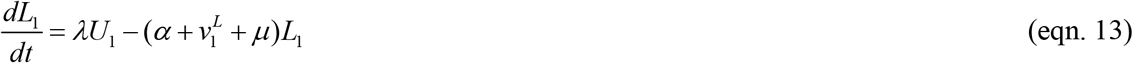

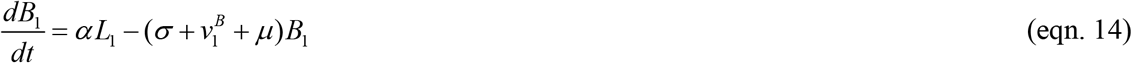

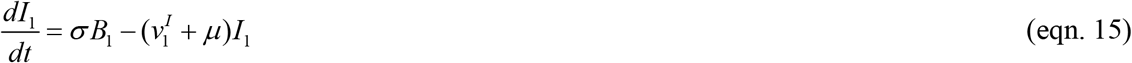

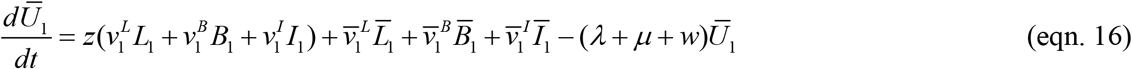

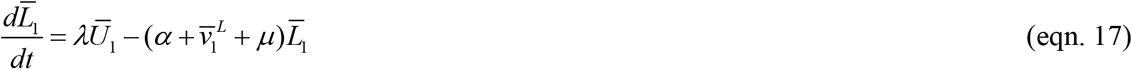

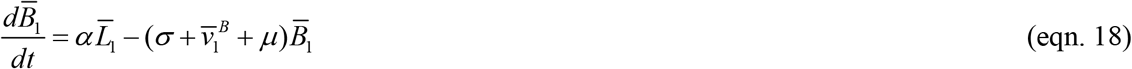

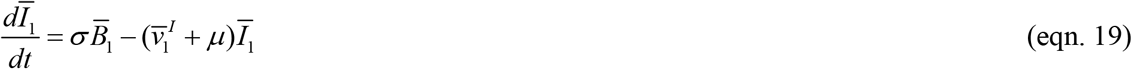

**Table S1.** Model parameters.

| Parameter | Description | Value | Source |
| --- | --- | --- | --- |
| $\theta$ | Proportion of population who participate in MDA | Variable | Varied to explore its impact |
| $p$ | The proportion of symptomatic infections that are treated amongst the portion of the population who do not participate in MDA. | Variable | Varied to explore its impact |
| $N$ | Total population | 1000 | [33]<sup>a</sup> |
| $\mu$ | Per-person death rate | 0.015 days<sup>-1</sup> | [34] |
| $\alpha$ | Per-person rate of transition from a liver-stage infection to a non-infectious blood stage infection | 1/5 days<sup>-1</sup> | [12] |
| $\sigma$ | Per-person rate of transition from an non-infectious to an infectious blood stage infection | 1/15 days<sup>-1</sup> | [12] |
| $w$ | The per-person rate that immunity is lost | 0.8/365 days<sup>-1</sup> (Supplementary Figure 2 blue line)<br>1.07/365 days<sup>-1</sup> (Otherwise) | [26] |
| $g$ | The ratio of the amplitude of the vertical shift of the transmission function ( $g = \text{amplitude}/\beta$ ) | 0.67 | [12] |
| $\eta$ | The ratio of infectiousness of immunologically experienced to immunologically inexperienced individuals. | 0.2 (Supplementary Figure 1 blue line)<br>1 (Otherwise) | [23] |
| $v_i^x$ | Per-person rate of recovery of infected individuals in MDA group $i$. The infection stage is denoted by $x \in \{L, B, I\}$ and the presence of therapy and immunological experience is denoted by the accent in accordance with the notation used for the state variables | $\bar{v}_0^I = \tilde{v}^I = 1 / 60 \text{ days}^{-1}$<br>$v_0^B = v_0^I = 1 \text{ days}^{-1}$ | [12]<br>[35, 36]<br>[35, 36]<br>[12]<br>[35, 36]<br>[12, 37] |
| | | $v_1^L = \bar{v}_1^L = \begin{cases} 1 \text{ days}^{-1} & (\text{days 1-3 of MDA}) \\ 0 \text{ days}^{-1} & (\text{days 4-25 of MDA}) \end{cases}$ $v_1^B = \bar{v}_1^B = \begin{cases} 1 \text{ days}^{-1} & (\text{days 1-3 of MDA}) \\ 1/3 \text{ days}^{-1} & (\text{days 4-25 of MDA}) \end{cases}$ $v_1^I = \bar{v}_1^I = \begin{cases} 1 \text{ days}^{-1} & (\text{days 1-3 of MDA}) \\ 1/21 \text{ days}^{-1} & (\text{days 4-25 of MDA}) \end{cases}$ | [35, 36]<br>[12, 37] |
| $z$ | Proportion of infections amongst immunologically inexperienced individuals that result in acquired immunity | 0.8 (Supplementary Figure 2 blue line)<br>0.5 Otherwise | [38] |
| $\beta$ | The transmission coefficient. This parameter scales the vertical shift of the transmission function. | 8.14 Figure 3a<br>8.72 Figure 4<br>Variable Otherwise | Fitted to give 30% malaria prevalence (with seasonality).<br>Fitted to give 30% malaria prevalence (without seasonality).<br>Varied to explore impact |

<sup>a</sup> This figure is based upon a pilot trial of MDA in four villages in Eastern Myanmar with a mean number of 810 in each village. For clarity, this number was rounded up to 1000.

**Table S2.**
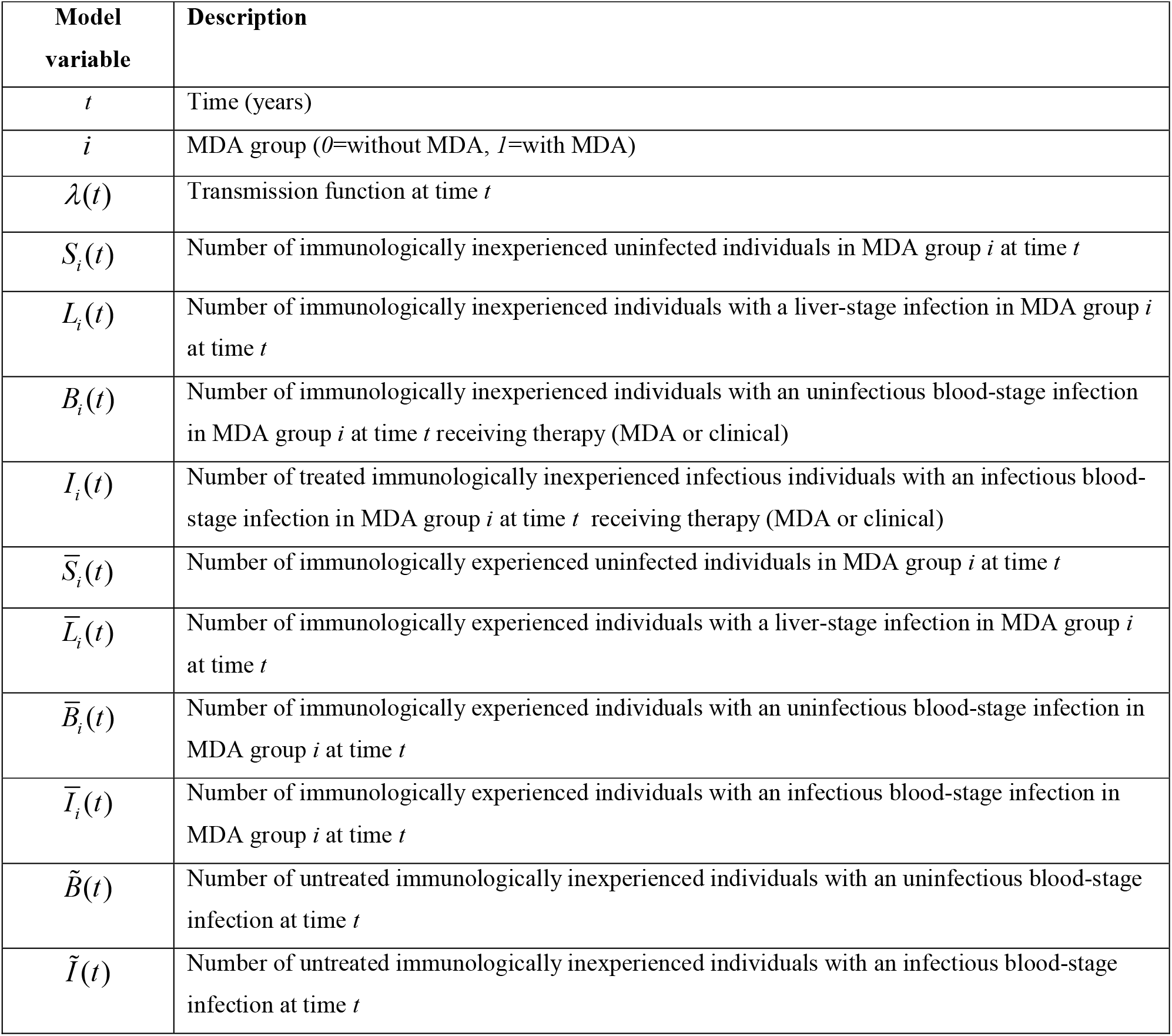
Model variables.

